# Modeling Prenatal Adversity/Advantage: Effects on Birth Weight

**DOI:** 10.1101/2021.12.16.21267938

**Authors:** Joan L. Luby, Deanna M. Barch, Barbara Warner, Cynthia Rogers, Chris Smyser, Regina Triplett, Jyoti Arora, Tara Smyser, Sarah K. England, George Slavich, Molly Stout, Phil Miller

**Affiliations:** Department of Psychiatry, Washington University School of Medicine; Department of Psychological & Brain Sciences, Washington University in St. Louis; Department of Pediatrics, Washington University School of Medicine; Department of Neurology, Washington University School of Medicine; Division of Biostatistics, Washington University School of Medicine; Department of Obstetrics and Gynecology, Center for Reproductive Health Sciences, Washington University School of Medicine; Department of Psychiatry and Biobehavioral Sciences, University of California, Los Angeles; Department of Obstetrics and Gynecology, University of Michigan

## Abstract

**Importance:** How maternal experiences of adversity/advantage during pregnancy impact the developing fetus remains unclear.

**Objective:** Using prospective data about experiences of adversity/advantage and other factors known to impact fetal developmental, we explored how these risk and protective factors relate to each other and impact infant birth weight by gestational age.

**Design:** A prospective study that collected data on of forms of social advantage/disadvantage, and psychological factors from pregnant women during each trimester of pregnancy that accounted for maternal medical and nutritional status. We aimed to determine the differential impact of social advantage/disadvantage and adversity and psychological factors on infant birthweight accounting for gestational age. Structural Equation Modeling (SEM) was used to investigate the relationship of these forms of adversity as latent constructs on infant outcome. The follow-up of children is ongoing as a part of the Early Life Adversity Biological Embedding and Risk for Developmental Precursors of Mental Disorders (eLABE). Data collection was conducted from 2017-2020.

**Setting:** An academic medical center.

**Participants:** Pregnant women who were participants in a study of preterm birth within the Prematurity Research Center at Washington University in St. Louis with negative drug screens (other than cannabis) and without known pregnancy complications or known fetal congenital problems, were invited for participation. N=395 mothers were included in the analysis and N=268 eligible subjects declined participation. N=399 singleton offspring were included.

**Main Outcome(s) and Measure(s):** Birthweight accounting for gestational age.

**Results:** The study included N=395 pregnant women and their N=399 singleton offspring. The Social Advantage latent factor significantly predicted the residual birthweight after accounting for gestational age (p=.006) representing a 2.57% increase in residual gestational age-adjusted birthweight for each one standard deviation increase in the Social Advantage. The only other significant predictor was pre-pregnancy BMI (p=.019) which was associated with increased birthweight by gestational age while the Psychosocial Stress factor was no longer significant when other factors were accounted for.

**Conclusions and Relevance:** Findings elucidate the significant effects of social adversity on the developing fetus and underscore the need to protect pregnant women in this risk group.

**Key Points:**

- Question: What are the differential effects of social adversity/advantage and psychosocial stress during pregnancy on fetal development evidenced by birthweight?
- Findings: Psychosocial adversity/advantage significantly impacted birthweight after accounting for other salient risk factors including maternal physical health.
- Meaning: Resources and interventions for pregnant women experiencing social adversity should become a public health priority for the protection of the developing fetus.

---

The theory of the developmental origins of health and disease (DOHaD) has focused scientific attention on the powerful impact of the intrauterine environment on neonatal health outcomes.^1^ Beyond the well-established effects of maternal physical health and nutritional status as well as drug and toxin exposure, more recent literature has emphasized the material importance of the maternal psychosocial environment on infant outcome, focusing on experiences of adversity and stress.^2^ Numerous studies have documented significant effects and have explored the mechanisms by which such psychosocial factors, conceptualized as “prenatal programming,” relate to a variety of infant outcomes. These studies emphasize the role for maternal psychological well-being. Specifically, the effect of stress and adversity versus that of advantage (material and social resources/support) on the developing fetus. Such effects have been conceptualized as “priming” for post-natal plasticity and adaptive mechanisms to prepare the developing infant for the expected external environment^3^.

A complex issue central to investigations of the effects of psychosocial adversity on health outcomes is the frequent co-occurrence of numerous forms of adversity, making it difficult to determine whether there are specific impacts of any particular type of adverse exposure. Researchers have attempted to decipher specific effects of forms of adversity by reporting on unique effects of deprivation and neglect as distinct from abuse and trauma.^4,5^ Others have focused on cumulative stress risk, and some have attempted to clarify the specific effects of different kinds of adversity such as exposure to violence or area deprivation index (e.g., median income, housing costs) as distinct from family income more generally.^6^ Additional work has examined the impact of maternal depression and related psychological stress during pregnancy on infant outcomes,^7^ but this work has not yet attempted to dissociate these factors from poverty or other forms of social disadvantage.^8^ Therefore, it remains unclear if there are effects of social adversity on prenatal programming that can be differentiated from the established effects of maternal depression during pregnancy. Based on this, there is a need for investigations that attempt to distinguish the effects of different forms of adversity in the prenatal period on fetal development. Understanding the relevant exposures and their collective effect on the developing fetus will be imperative to inform targeted prevention.

In an ongoing longitudinal study, we collected data from pregnant women during each trimester of pregnancy to determine the impact of a variety of forms of social advantage/disadvantage and adversity and psychological factors such as depression, psychosocial stress, and stressful and traumatic life events on neonatal outcomes. Here we focus on birthweight accounting for gestational age as a key early neonatal outcome known to be associated with later health trajectories, and when adjusted for brain volume, with childhood IQ.^9^ Birthweight is one of the very first indicators of later health and developmental outcomes with extremes of birthweight, such as small for gestational age (SGA; <10^th^ percentile at birth) and large for gestational age (LGA; >90^th^ percentile at birth), established as sensitive markers of cardiometabolic and neurodevelopmental risk into adulthood.^10,11^ While DOHaD originally emphasized poor long term outcomes associated with these extremes of birthweight, data also suggest a relationship between birthweight within the normative spectrum to later childhood cognitive outcomes.^12-14^ Further, one twin birth cohort has shown incremental weight differences across the normative birthweight spectrum to be related to adult cognitive, educational, and earning achievements.^15^ Thus, while the majority of the literature on prenatal medical and psychosocial determinants has mainly focused on extremes,^16-18^ further work examining these effects along a normative continuum to adverse later life health outcomes are needed.^19^ This study aims to fill this gap by examining the relationships of maternal medical, psychosocial and psychological (i.e., stress, anxiety, depression), and social determinants of health (i.e., income to needs, nutrition, neighborhood, etc.) to the normative spectrum of birthweight given that this metric is a key predictor of later health trajectories.

With the aim of investigating and disentangling the high levels of correlation between various forms of adversity, Structural Equation Modeling (SEM) was used to test the differential relationships of forms of adversity to birthweight accounting for gestational age and sex. SEM is particularly useful for examination of complex relationships between measured variables that may be inter-related given its ability to identify and measure dissociable latent constructs. We used a range of measured variables to define two latent constructs, Maternal Social Advantage and Maternal Psychosocial Stress. Along with maternal medical risk factors, the contribution of these constructs, along with their individual components, to birthweight was examined. Birthweight was used because of its importance to long term health outcomes, and to assess its relationship to Social Advantage and Psychosocial Stress before assessing relationships of these adversity constructs to direct measures of neurodevelopment including brain structure (see Tripett et al.,) and function, as well as neurobehavioral assessments at birth. We examined whether there were dissociations between the relationships of social disadvantage, indexed by poverty, area deprivation, low maternal education, and poor nutrition, versus psychosocial disadvantage, indexed by maternal depression, discrimination, and experiences of stress and trauma, in predicting lower birthweight even when accounting for maternal health parameters including medical risk, body mass index, and maternal age. Within this cohort, race was highly correlated to Social Advantage Factor indices, offering no additional improvement to the model after other variables (including discrimination) were accounted for and thus it was not included as a variable.

## Methods

The current study, Early Life Adversity Biological Embedding and Risk for Developmental Precursors of Mental Disorders (eLABE), is a multi-wave, multi-method NIMH-funded study designed to investigate the mechanisms by which prenatal and early life adversity impact infant neurodevelopment. Pregnant women who were participants in a large-scale study of preterm birth within the Prematurity Research Center at Washington University in St. Louis with negative drug screens (other than cannabis) and without known pregnancy complications or known fetal congenital problems, were invited for eLABE participation. The study recruited N=395 women during pregnancy (N=268 eligible subjects declined participation) and their N=399 singleton offspring (N=4 mothers had 2 singleton births during the recruitment period). Out of those originally invited and interested in participation, N=26 were deemed ineligible (N=13 screened out prior to consent and N=13 consented subjects were deemed ineligible due to later discovery of substance abuse or the finding of a congenital anomalies). Women facing social disadvantage were over-sampled by increased recruitment from a clinic serving low-income women. The sample was also enriched for preterm infants with N=51 born preterm (<37 weeks gestation). Of the 399 pregnancies, 50 reported tobacco use during pregnancy, and 49 reported cannabis use (of which 20 reported both). See *Supplemental Materials* for analyses that controlled for use of cannabis and tobacco.

As described, maternal depression, experiences of stress, as well as demographic information including insurance, education, address, and household composition were obtained from participants at each trimester during pregnancy. Birthweight and gestational age of neonates, as well as maternal dietary and medical history, were obtained from self-reported surveys and medical records. Mothers and their newborns were invited for an assessment shortly after birth which included neonatal MRI during which mothers completed a comprehensive measure of life stress and trauma (current and past) and discrimination. Measures are detailed in Table S1.

We hypothesized that we could dissociate and find significant independent relationships of **Maternal Social Advantage** versus **Maternal Psychosocial Stress** to birthweight. To test this hypothesis, we first conducted a confirmatory factor analysis for *a priori* hypotheses about which indicators loaded on these two factors. This model incorporates latent variables for these two domains which are assumed to be measured without error. Each is indexed by a group of relevant measured variables thought to be associated with the latent variable. The degree of association between each measured variable and the latent variable is a function of both measurement reliability and concept overlap. A multiple regression SEM model then looked at the independent contribution of each of these latent variables and **Maternal Medical Factors** in predicting birthweight adjusted for gestational age.

### Maternal Social Advantage

<u>Income to Needs ratio</u> (I/N) was measured at each trimester. The I/N ratio utilizes self-reported family income and household size compared to federal poverty thresholds, with a ratio of 1.0 being at the poverty line. <u>Insurance status</u> was collected at the time of enrollment through a medical record review and was verified in the third trimester or at delivery; Mother’s <u>highest level of education</u> was self-reported at the time of enrollment. <u>Area Deprivation Index</u> (ADI) is a geotracking measure was used to rank neighborhoods by socioeconomic disadvantage compared to the national average based on census block data, including factors for the domains of income, education, employment, and housing quality^20,21^. <u>Maternal nutrition</u> was assessed using the Healthy Eating Index (HEI) during the third trimester or at delivery. This is a validated dietary assessment tool available through the National Institutes of Health used to measure diet quality based on U.S Dietary Guidelines for Americans.^22,23^ Dietary information for HEI calculation was obtained using the Diet History Questionnaire (DHQII).^24,25^

### Maternal Psychosocial Stress

In each trimester, mothers completed the Edinburgh Postnatal Depression Scale (EPDS)^26^ and Perceived Stress scale (PSS)^27^. The STRAIN^28^, a comprehensive measure of lifetime stressful and traumatic life events, was collected at the time of neonatal scan (N=255) or at a follow-up exam (N=108); no differences in STRAIN scores based on time of administration were found. Experiences of discrimination based on race were assessed using the Everyday Discrimination Scale^29^ measured at neonatal scan.

### Maternal Medical Factors

To control for maternal medical risks that might be confounded with social or psychological disadvantage, we assessed maternal age at delivery and pre-pregnancy body mass index from first prenatal visit based on self-reported pre-pregnancy weight and height. In addition, a Maternal Medical Risk Score (MMR), containing pre-existing and pregnancy related medical conditions, was computed using a validated measure of maternal medical comorbidities extracted from the medical record that accounts for 22 medical conditions weighted by severity.^30,31^ Because of the potential for a direct effect of maternal nutrition on birthweight, a direct link was included for HEI.

### Child Variables

Birthweight was collected from the medical record at the time of delivery. In addition, we used gestational age as determined by best obstetric estimate using last menstrual period or earliest ultrasound dating available at birth as a covariate.

### Statistical Methods

Confirmatory factor analysis (as distinct from exploratory factor analysis) prespecifies the number of factors and which variables will load on which factor. It was undertaken using the MPlus software to validate our grouping of prenatal adversity variables into a **Social Advantage** latent factor reflecting indicators of social advantage/disadvantage (I/N, ADI, Insurance Status, Maternal Education, and Maternal Nutrition) and a **Psychosocial Stress** factor (depression, perceived stress, discrimination, and lifetime measures of trauma and life events) as indicated in Table 1. Education and Insurance were specified as categorical variables within the Social Advantage factor. The variances of the Social Advantage and Psychosocial factors were set at 1.0. The model is diagramed in Figure 1. In fitting the model, we examined the residual correlations which remained unexplained by the prespecified model and added covariances where necessary. In particular, we examined whether any of the variables associated with the latent factors also had relationships to birthweight which were not accounted for by the latent variable, which would indicate a direct effect for that variable in addition to that contributed by the latent variable. The estimation of the model was with the MLR maximum likelihood method which allows for estimation even in the setting of missing data. The STDY standardization of the effects was used. To adjust birthweight for gestational age, we fit a loess regression model using the default parameters in Proc LOESS of SAS. The residuals from this regression were then used as the dependent measure assessing the degree that the weight of the infant was heavier or lighter than average for gestational age. Because of observed heteroscedasticity and a lower degree of correlation between birthweight and gestational age with raw birthweight in grams (r=.69) versus with log_10_ transformed birthweight (r=0.77), a log_10_ transformation of birthweight was used. Because of reports of differing imprinting by the sex of the fetus,^32,33^ we also tested interactions of sex with each of the latent variables. All computations were done with MPlus version 8.4 or SAS 9.4.

**Table 1.**
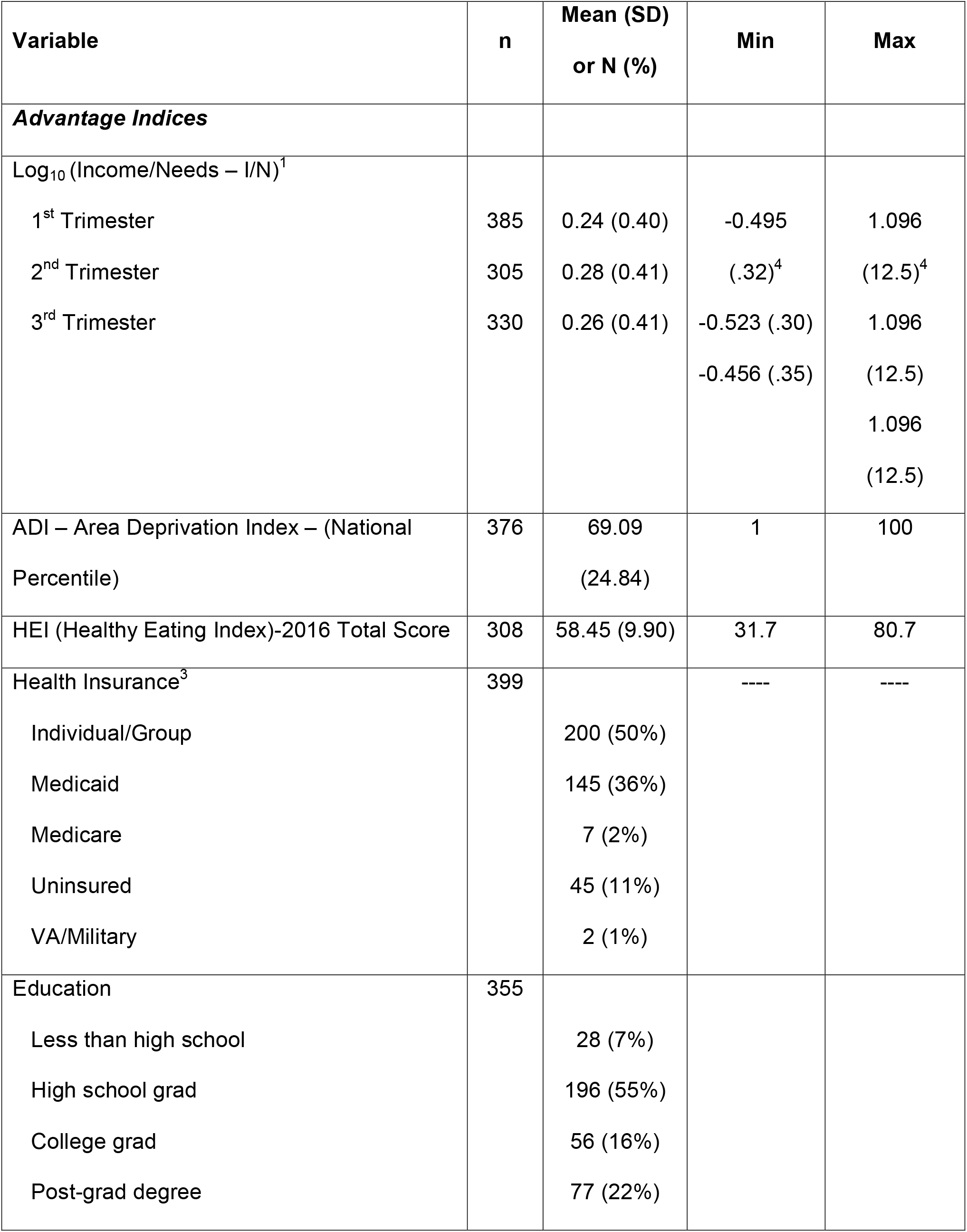
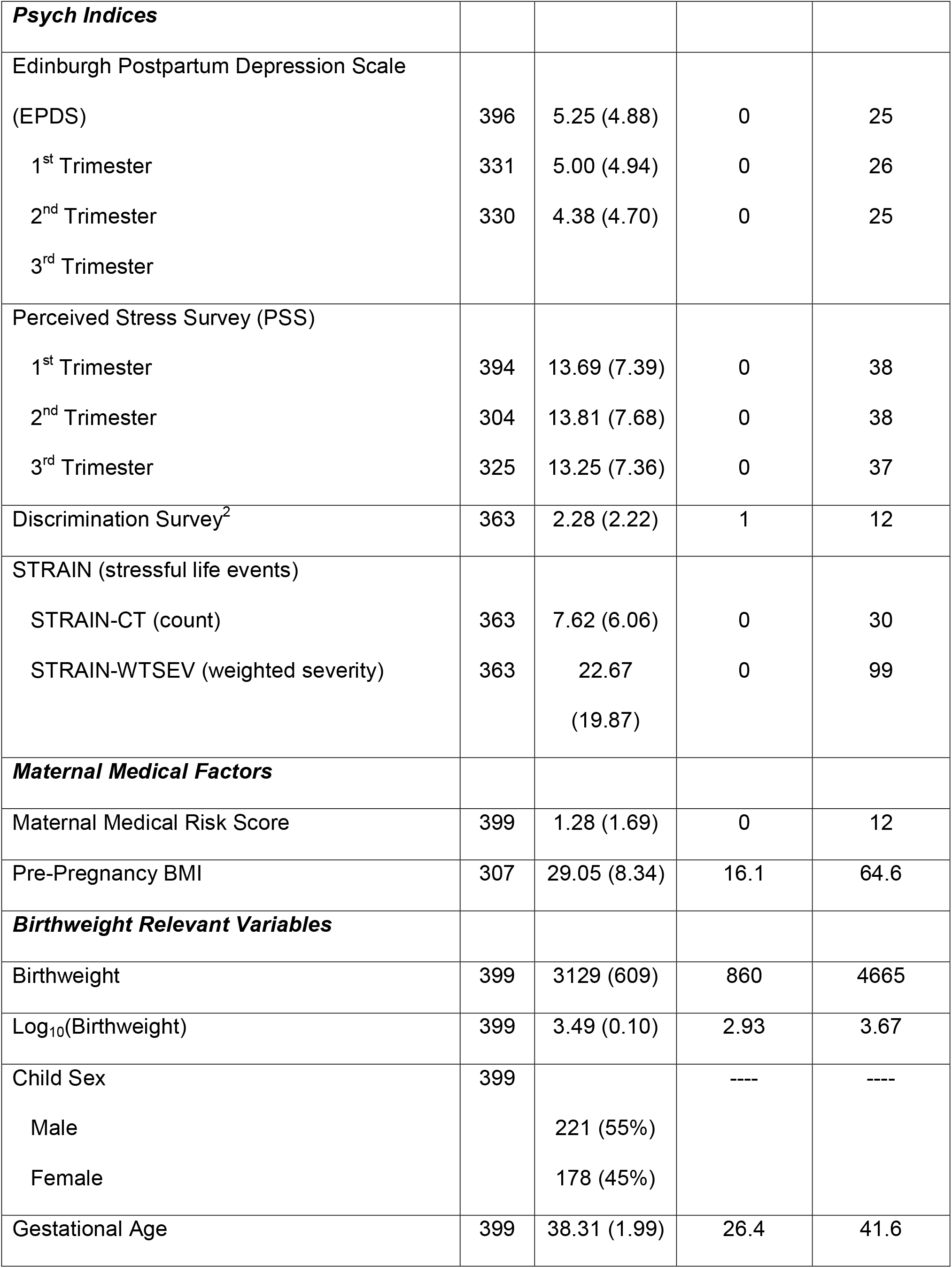
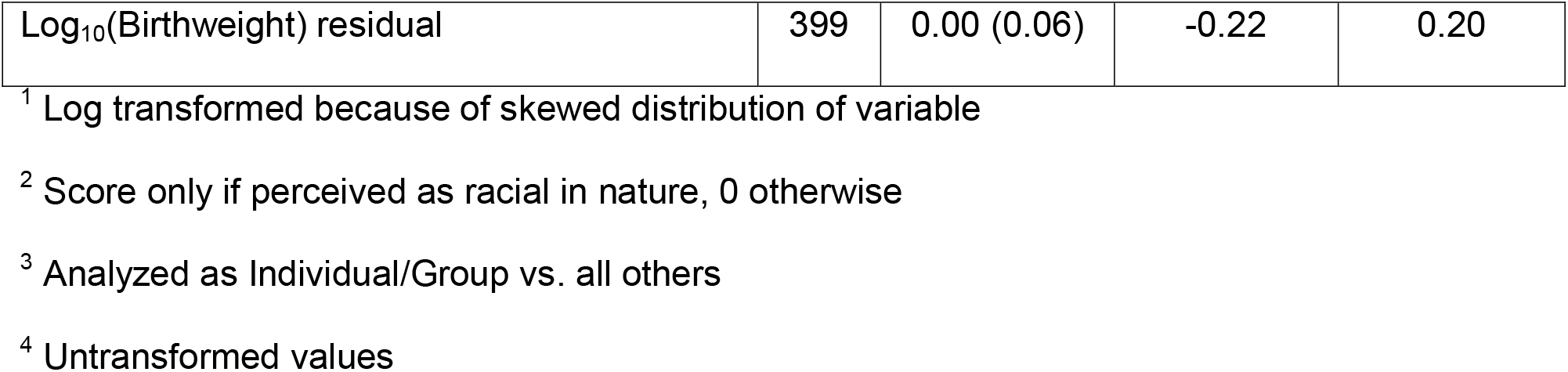
Measured Maternal and Child Variables in SEM Model.

| Variable | n | Mean (SD)<br>or N (%) | Min | Max |
| --- | --- | --- | --- | --- |
| ***Advantage Indices*** |  |  |  |  |
| $\text{Log}_{10} (\text{Income/Needs} - \text{I/N})^1$ | | | | |
| 1<sup>st</sup> Trimester | 385 | 0.24 (0.40) | -0.495 | 1.096 |
| 2<sup>nd</sup> Trimester | 305 | 0.28 (0.41) | (.32)<sup>4</sup> | (12.5)<sup>4</sup> |
| 3<sup>rd</sup> Trimester | 330 | 0.26 (0.41) | -0.523 (.30)<br>-0.456 (.35) | 1.096<br>(12.5)<br>1.096<br>(12.5) |
| ADI – Area Deprivation Index – (National Percentile) | 376 | 69.09<br>(24.84) | 1 | 100 |
| HEI (Healthy Eating Index)-2016 Total Score | 308 | 58.45 (9.90) | 31.7 | 80.7 |
| Health Insurance<sup>3</sup> | 399 |  | ---- | ---- |
| Individual/Group |  | 200 (50%) |  |  |
| Medicaid |  | 145 (36%) |  |  |
| Medicare |  | 7 (2%) |  |  |
| Uninsured |  | 45 (11%) |  |  |
| VA/Military |  | 2 (1%) |  |  |
| Education | 355 |  |  |  |
| Less than high school |  | 28 (7%) |  |  |
| High school grad |  | 196 (55%) |  |  |
| College grad |  | 56 (16%) |  |  |
| Post-grad degree |  | 77 (22%) |  |  |
| ***Psych Indices*** |  |  |  |  |
| Edinburgh Postpartum Depression Scale (EPDS) | 396 | 5.25 (4.88) | 0 | 25 |
| 1<sup>st</sup> Trimester | 331 | 5.00 (4.94) | 0 | 26 |
| 2<sup>nd</sup> Trimester | 330 | 4.38 (4.70) | 0 | 25 |
| 3<sup>rd</sup> Trimester |  |  |  |  |
| Perceived Stress Survey (PSS) |  |  |  |  |
| 1<sup>st</sup> Trimester | 394 | 13.69 (7.39) | 0 | 38 |
| 2<sup>nd</sup> Trimester | 304 | 13.81 (7.68) | 0 | 38 |
| 3<sup>rd</sup> Trimester | 325 | 13.25 (7.36) | 0 | 37 |
| Discrimination Survey<sup>2</sup> | 363 | 2.28 (2.22) | 1 | 12 |
| STRAIN (stressful life events) |  |  |  |  |
| STRAIN-CT (count) | 363 | 7.62 (6.06) | 0 | 30 |
| STRAIN-WTSEV (weighted severity) | 363 | 22.67<br>(19.87) | 0 | 99 |
| ***Maternal Medical Factors*** |  |  |  |  |
| Maternal Medical Risk Score | 399 | 1.28 (1.69) | 0 | 12 |
| Pre-Pregnancy BMI | 307 | 29.05 (8.34) | 16.1 | 64.6 |
| ***Birthweight Relevant Variables*** |  |  |  |  |
| Birthweight | 399 | 3129 (609) | 860 | 4665 |
| Log<sub>10</sub>(Birthweight) | 399 | 3.49 (0.10) | 2.93 | 3.67 |
| Child Sex | 399 |  | ---- | ---- |
| Male |  | 221 (55%) |  |  |
| Female |  | 178 (45%) |  |  |
| Gestational Age | 399 | 38.31 (1.99) | 26.4 | 41.6 |
| Log<sub>10</sub>(Birthweight) residual | 399 | 0.00 (0.06) | -0.22 | 0.20 |

<sup>1</sup> Log transformed because of skewed distribution of variable
<sup>2</sup> Score only if perceived as racial in nature, 0 otherwise
<sup>3</sup> Analyzed as Individual/Group vs. all others
<sup>4</sup> Untransformed values

**Figure 1.**
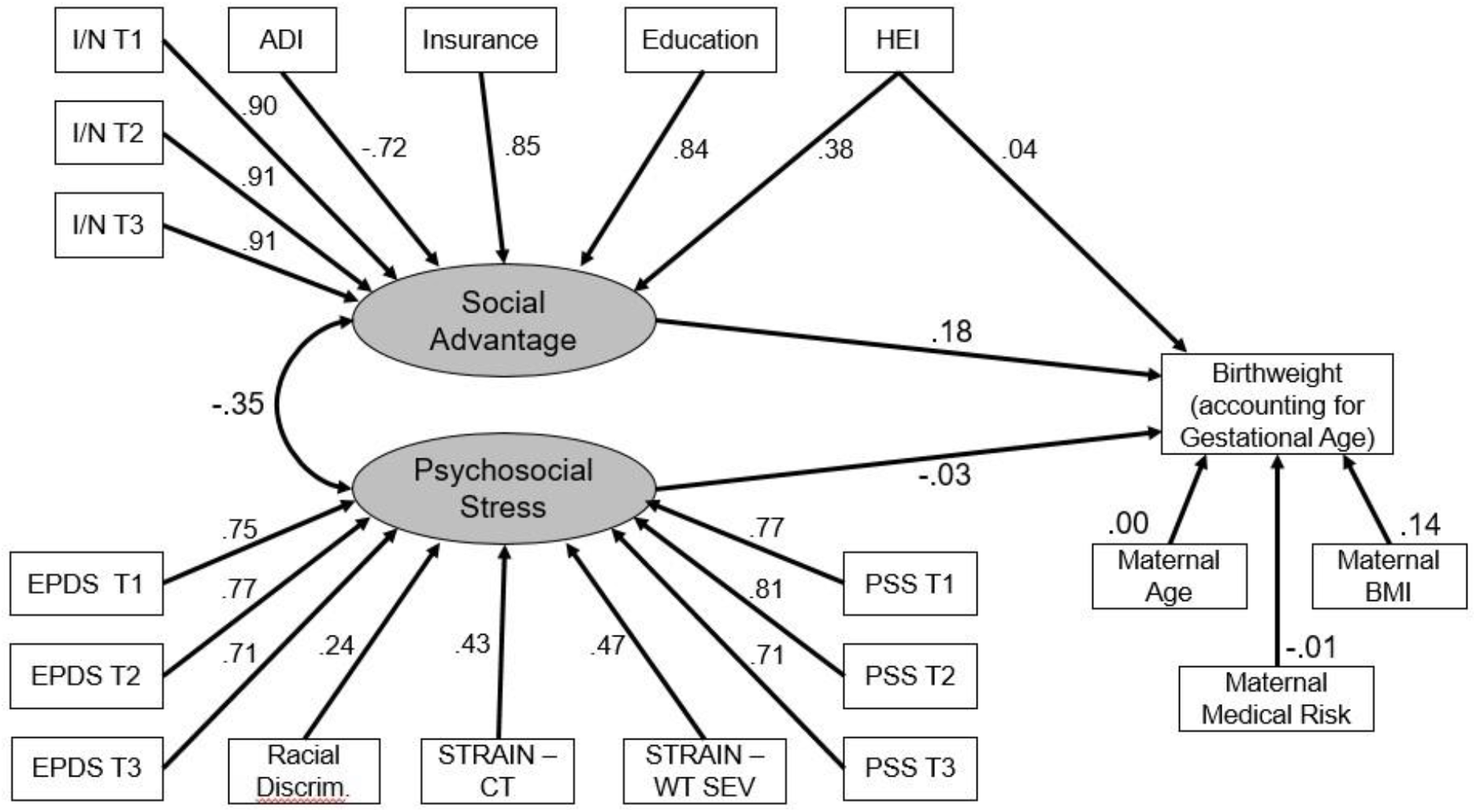
Structural Equation Model (note that some covariances are not included for clarity; see Table 2).

## Results

Table 1 presents means and standard deviations for each of the individual measures including the Social Advantage and Psychosocial Stress factors, as well as for the maternal medical risk and child outcome measures. While the cohort is weighted toward socially disadvantaged mothers as described in the Methods, a broad range across social and psychological variables exist, with 38% of the population having a college degree or higher, over 50% of the population with private insurance, and I/N values extending up to 12 times above the poverty level.

The relationships of each of the measured variable to the latent variables are shown in Figure 1 and Table 2. The results of this confirmatory factor produced fit indices that indicated a good level of fit (RMSEA=0.042, SRMR=0.055, CFI/TU=0.995/0.946). Importantly, there were low levels of correlations of the measured variables for one factor (e.g., Social Advantage) with the other factor. We then compared this two-factor model to a one-factor model and none of the fit indices were in an acceptable range (RMSEA=0.14, SRMR=0.15, CFI/TLI=0.512/0.408). A three-factor model had two fit indicators in an acceptable range (RMSEA=0.080, SMSR=0.080), but CFI/TLI was 0.844/0.808 and not in an acceptable range. In addition, the loadings on the third factor were all low. Notably, for the two-factor model, there did not appear to be differential relationships of indicators of either the Social Advantage or Psychosocial factors as a function of trimester, as the loadings were approximately equal across trimesters (Table 2).

**Table 2:**
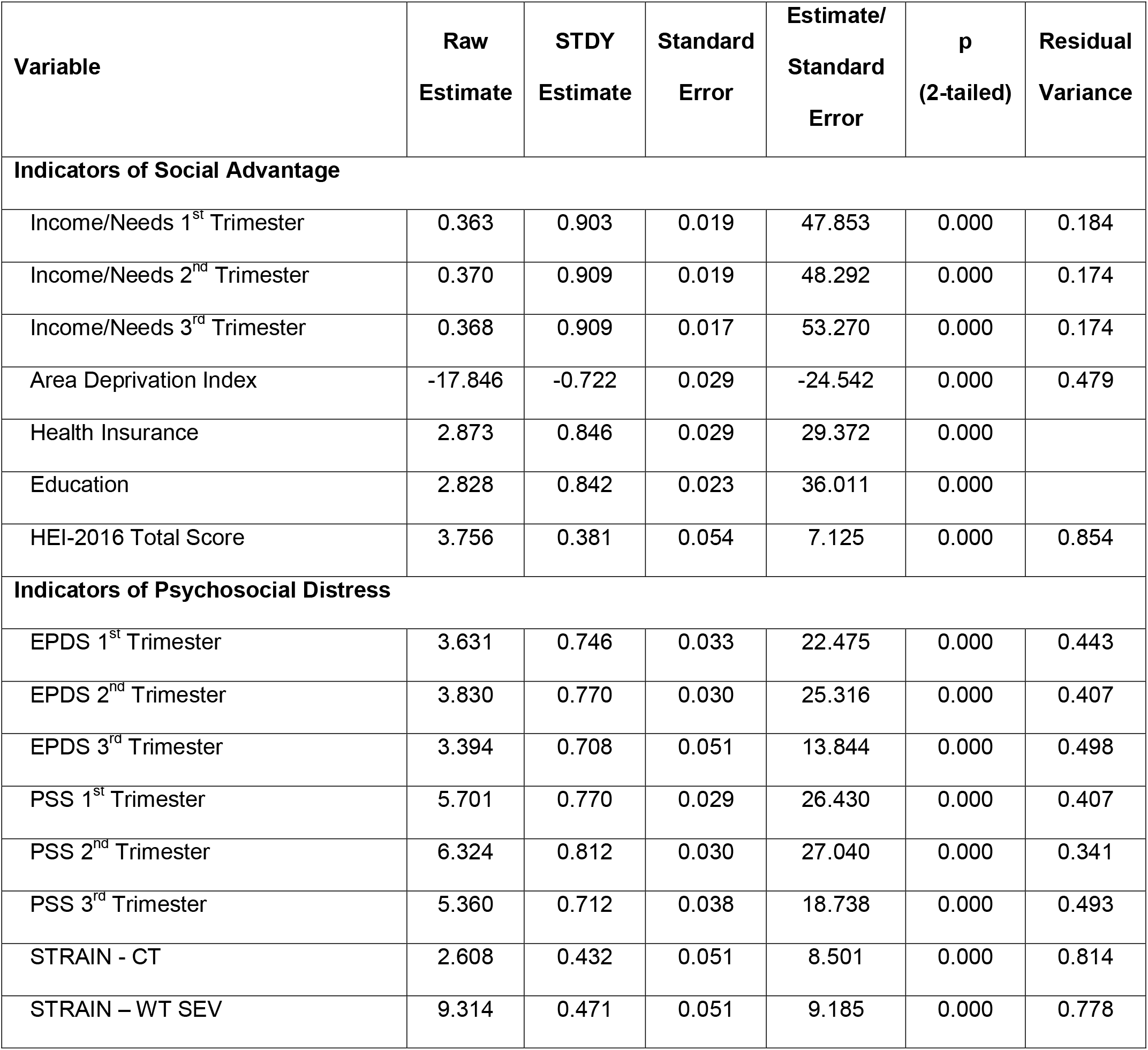
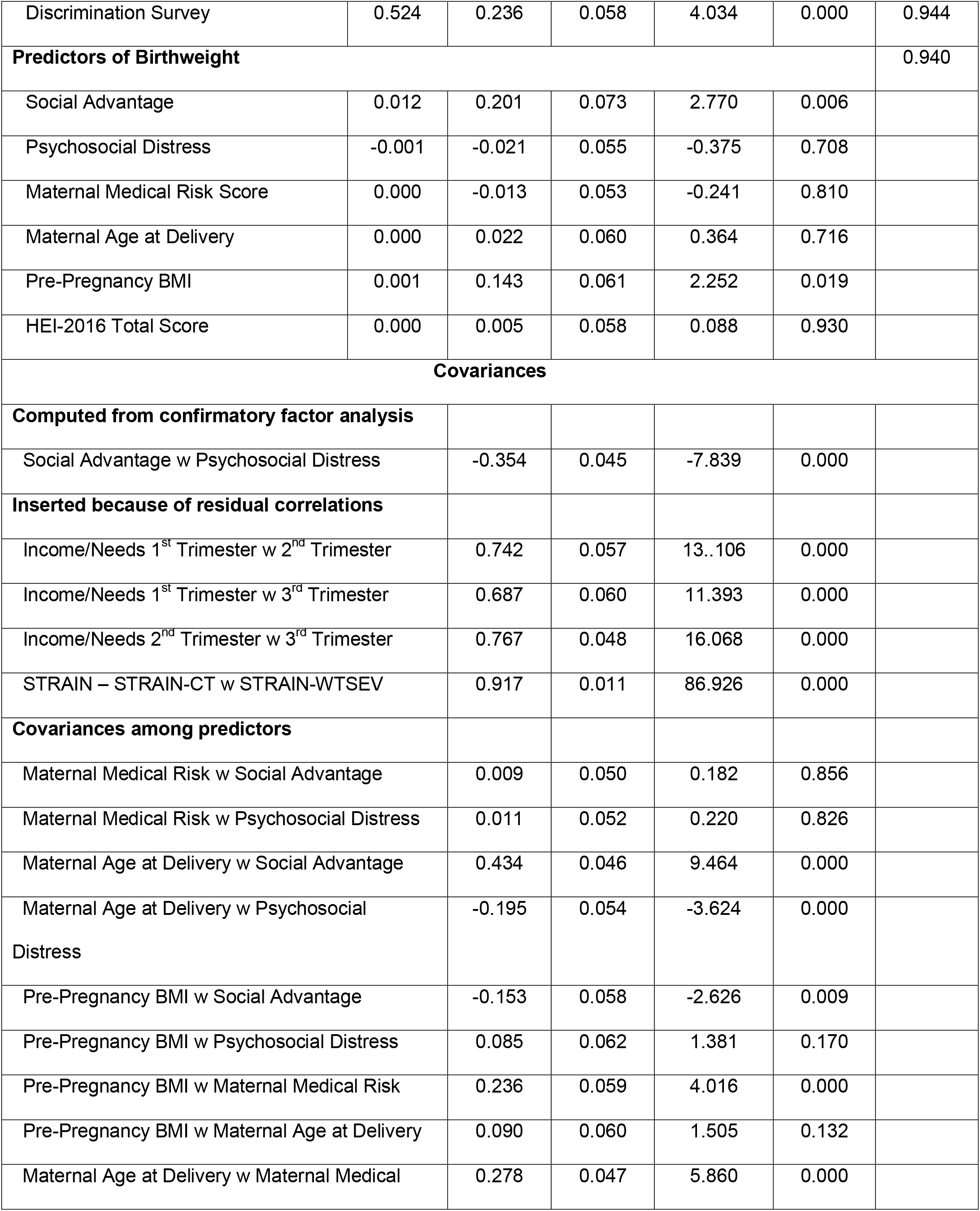
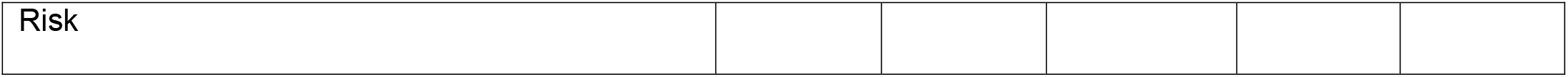
Variables Included in SEM Model.

| Variable | Raw<br>Estimate | STDY<br>Estimate | Standard<br>Error | Estimate/<br>Standard<br>Error | p<br>(2-tailed) | Residual<br>Variance |
| --- | --- | --- | --- | --- | --- | --- |
| **Indicators of Social Advantage** |  |  |  |  |  |  |
| Income/Needs 1<sup>st</sup> Trimester | 0.363 | 0.903 | 0.019 | 47.853 | 0.000 | 0.184 |
| Income/Needs 2<sup>nd</sup> Trimester | 0.370 | 0.909 | 0.019 | 48.292 | 0.000 | 0.174 |
| Income/Needs 3<sup>rd</sup> Trimester | 0.368 | 0.909 | 0.017 | 53.270 | 0.000 | 0.174 |
| Area Deprivation Index | -17.846 | -0.722 | 0.029 | -24.542 | 0.000 | 0.479 |
| Health Insurance | 2.873 | 0.846 | 0.029 | 29.372 | 0.000 |  |
| Education | 2.828 | 0.842 | 0.023 | 36.011 | 0.000 |  |
| HEI-2016 Total Score | 3.756 | 0.381 | 0.054 | 7.125 | 0.000 | 0.854 |
| **Indicators of Psychosocial Distress** |  |  |  |  |  |  |
| EPDS 1<sup>st</sup> Trimester | 3.631 | 0.746 | 0.033 | 22.475 | 0.000 | 0.443 |
| EPDS 2<sup>nd</sup> Trimester | 3.830 | 0.770 | 0.030 | 25.316 | 0.000 | 0.407 |
| EPDS 3<sup>rd</sup> Trimester | 3.394 | 0.708 | 0.051 | 13.844 | 0.000 | 0.498 |
| PSS 1<sup>st</sup> Trimester | 5.701 | 0.770 | 0.029 | 26.430 | 0.000 | 0.407 |
| PSS 2<sup>nd</sup> Trimester | 6.324 | 0.812 | 0.030 | 27.040 | 0.000 | 0.341 |
| PSS 3<sup>rd</sup> Trimester | 5.360 | 0.712 | 0.038 | 18.738 | 0.000 | 0.493 |
| STRAIN - CT | 2.608 | 0.432 | 0.051 | 8.501 | 0.000 | 0.814 |
| STRAIN – WT SEV | 9.314 | 0.471 | 0.051 | 9.185 | 0.000 | 0.778 |
| Discrimination Survey | 0.524 | 0.236 | 0.058 | 4.034 | 0.000 | 0.944 |
| **Predictors of Birthweight** |  |  |  |  |  | 0.940 |
| Social Advantage | 0.012 | 0.201 | 0.073 | 2.770 | 0.006 |  |
| Psychosocial Distress | -0.001 | -0.021 | 0.055 | -0.375 | 0.708 |  |
| Maternal Medical Risk Score | 0.000 | -0.013 | 0.053 | -0.241 | 0.810 |  |
| Maternal Age at Delivery | 0.000 | 0.022 | 0.060 | 0.364 | 0.716 |  |
| Pre-Pregnancy BMI | 0.001 | 0.143 | 0.061 | 2.252 | 0.019 |  |
| HEI-2016 Total Score | 0.000 | 0.005 | 0.058 | 0.088 | 0.930 |  |
| **Covariances** |  |  |  |  |  |  |
| **Computed from confirmatory factor analysis** |  |  |  |  |  |  |
| Social Advantage w Psychosocial Distress |  | -0.354 | 0.045 | -7.839 | 0.000 |  |
| **Inserted because of residual correlations** |  |  |  |  |  |  |
| Income/Needs 1<sup>st</sup> Trimester w 2<sup>nd</sup> Trimester |  | 0.742 | 0.057 | 13..106 | 0.000 |  |
| Income/Needs 1<sup>st</sup> Trimester w 3<sup>rd</sup> Trimester |  | 0.687 | 0.060 | 11.393 | 0.000 |  |
| Income/Needs 2<sup>nd</sup> Trimester w 3<sup>rd</sup> Trimester |  | 0.767 | 0.048 | 16.068 | 0.000 |  |
| STRAIN – STRAIN-CT w STRAIN-WTSEV |  | 0.917 | 0.011 | 86.926 | 0.000 |  |
| **Covariances among predictors** |  |  |  |  |  |  |
| Maternal Medical Risk w Social Advantage |  | 0.009 | 0.050 | 0.182 | 0.856 |  |
| Maternal Medical Risk w Psychosocial Distress |  | 0.011 | 0.052 | 0.220 | 0.826 |  |
| Maternal Age at Delivery w Social Advantage |  | 0.434 | 0.046 | 9.464 | 0.000 |  |
| Maternal Age at Delivery w Psychosocial Distress |  | -0.195 | 0.054 | -3.624 | 0.000 |  |
| Pre-Pregnancy BMI w Social Advantage |  | -0.153 | 0.058 | -2.626 | 0.009 |  |
| Pre-Pregnancy BMI w Psychosocial Distress |  | 0.085 | 0.062 | 1.381 | 0.170 |  |
| Pre-Pregnancy BMI w Maternal Medical Risk |  | 0.236 | 0.059 | 4.016 | 0.000 |  |
| Pre-Pregnancy BMI w Maternal Age at Delivery |  | 0.090 | 0.060 | 1.505 | 0.132 |  |
| Maternal Age at Delivery w Maternal Medical |  | 0.278 | 0.047 | 5.860 | 0.000 |  |

|  |
| --- |
| Risk |

The raw and standardized estimated effects for the whole group model are shown in Figure 1 and Table 2. The Social Advantage latent factor significantly predicted the residual birthweight after accounting for gestational age (p=.006). In examining the raw coefficient (0.012), this corresponds to a 2.57% increase in residual gestational age-adjusted birthweight for each one standard deviation increase in the Social Advantage factor (see Figure 2). In contrast, the Psychosocial Stress factor did not significantly predict residual birthweight. The only other significant predictor was a positive association of pre-pregnancy BMI (p=.019) with increased birthweight after accounting for gestational age. Social disadvantage continued to show a significantly direct effect in predicting residual birthweight even in an SEM model that accounted for cannabis and tobacco use, both of which were correlated with social disadvantage (See *Supplemental Materials* and Table S2).

**Figure 2.**
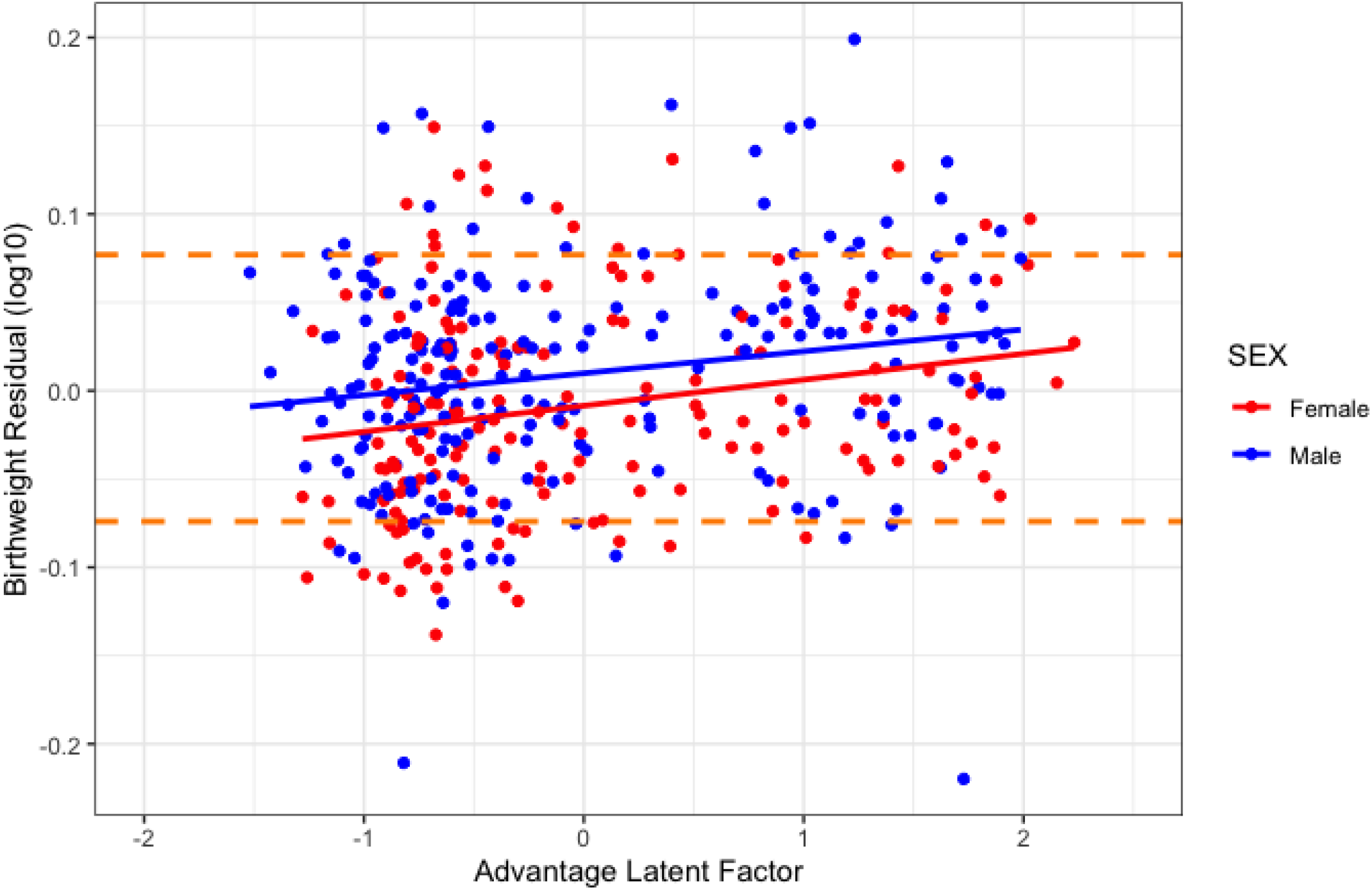
Estimated Advantage Latent Factor vs Birthweight Residual. Horizontal orange lines are 90^th^ and 10^th^ percentiles for this cohort.

We next tested interactions of sex with each of the latent factors. There were no significant interactions of sex with either the Social Advantage (p=.123) or Psychosocial (p=.457) factor in predicting birthweight.

## Discussion

The findings from these analyses demonstrate the central relationship of Social Advantage, a latent factor that includes income to needs, insurance status, education, area deprivation, and maternal nutrition, to birthweight adjusted for gestational age. While previous studies have focused on how prenatal economic or psychological stressors contribute specifically to low birth weight outcomes,^16,17^ few have examined their contribution when combined, or across the continuum of birthweight (see Figure 2). Notably, the Social Advantage score significantly predicted an increase in birthweight relative to gestational age by an increase in 2.57% for every 1 standard deviation in Advantage. To concretize this, for an infant born at the average gestational age in our sample (38.3 weeks), each standard deviation difference in Social Advantage translates to a difference of ∼90 grams in birthweight. The potential clinical significance of this might be contextualized by prior findings reporting that every 100 grams of birthweight change was associated with later IQ difference of .49.^9^ Further, the examination of both latent factors simultaneously allowed us to distinguish the importance of Social Advantage factors as distinct from Psychosocial Stress in contributing to birthweight outcomes. This Social Advantage factor predicted this key neonatal metric even when other critical markers of health and factors well-known to predict birthweight are accounted for in the model. The only other significant predictor was maternal BMI. This combination of results elucidates the critical role that Social Advantage experienced during pregnancy plays in fetal outcome.

Birthweight is a critical early indicator of risk for later cardiometabolic and neurodevelopmental risk into adulthood,^10,11^ as well as child IQ and adult cognitive, educational, and earning achievements.^9,15^ Our findings support that the relationship between maternal adversity and birthweight is relatively continuous, and shows significant associations extending into the normative range. Such findings underscore the need for more focused public health attention to these social disadvantage factors in pregnancy that have clear impact on neonatal birth outcomes, with lower birthweight a harbinger of poor physical, cognitive, and emotional health later in development. Additional analyses detailed in Triplett, et al. report on the significant relationships of these factors to measures of structural brain development at birth.

This SEM model elucidated how different indicators of adversity cohere to form factors (i.e., Social Advantage and Psychosocial Stress), as well as disentangled the relationships of different components of adversity to birthweight outcomes. The use of this modelling approach revealed several additional findings. First, it was notable that we did not see differential effects of exposures by trimester. Some specific effects of risk exposures during each trimester of pregnancy have been described in the extant literature.^34,35^ However, in our sample, the indicators of social advantage were stable across trimesters, which may have precluded us from identifying trimester specific effects. It is possible that samples with more variability in metrics of social advantage or psychosocial distress across pregnancy within mothers may reveal evidence for trimester specific effects. Further, it is also possible that trimester effects will become evident in later infant outcomes as “sleeper effects”. Second, it is notable that we did not find relationships of maternal psychosocial distress as indexed by depression, perceived distress, experience of discrimination, and life events once indicators of social advantage/disadvantage were included in the model. This finding contrasts with previous work reporting relationships of maternal depression and stress to neonatal outcomes.^8^ However, many of these prior studies did not account for socioeconomic indicators that are often co-occurring with psychosocial distress. Despite similar levels of adversity between our sample and several reported in the literature, our use of SEM revealed that at least in terms of birthweight, the socioeconomic factors played a more central role. It is possible that stronger relationships to psychosocial distress will emerge when other outcomes are examined, including brain development, behavior, emotional, and physical health. In addition, our sample utilized self-report measures of depression rather than diagnostic measures, and rates were somewhat lower than those in samples of clinically depressed pregnant women, which might have led to smaller than expected effects in this domain. Third, we were not able to investigate the role of race in this analysis due to the high co-linearity between race and SES in this sample. While race and class are highly linked in US samples, it is clear that race and class also intersect in important ways that relate to experiences of discrimination and social rejection, factors which will be an important area of focus in this sample during early development.^36,37^

These data highlight the central importance of maternal experiences of social advantage and disadvantage during pregnancy in predicting a key early outcome for offspring – birthweight. Given that birthweight is an infant outcome known to predict numerous later health trajectories, these findings further validate the principle that the social determinants of health are initiated during pregnancy. Further, this model provides an organizational framework to inform how the effects of adversity/advantage might be accounted for in studies of prenatal programming. Future studies from these data will utilize factors derived from this model to explore the relationships of social advantage and psychosocial distress to neonatal brain outcomes at birth, and to behavioral and brain outcomes over the course of early childhood development. However, current findings point to the importance of these forms of adversity/advantage and their differential contributions to neonatal birth outcomes suggesting they should be key health prevention targets in prenatal health care.

## Data Availability

All data produced in the present study are contained in the manuscript.

