## Supplement 1 for "Modeling Prenatal Adversity/Advantage: Effects on Birth Weight"

**Supplemental Materials**

**Supplemental Analyses Addressing Cannabis and Tobacco**

Cannabis and tobacco use can be more frequent among women who are experiencing social disadvantage and/or psychological distress and both cannabis and tobacco use have been associated with reduced birthweight.^36^ Thus, cannabis and/or tobacco use might be a confound in interpreting the relationships between social advantage and birthweight. Of the 399 women in this sample, 29 women reported using 1-5 cigarettes per day, and 21 reported 6 or more cigarettes per day, which was coded as 0/1/2 for analyses. Twenty women reported using cannabis at least one time weekly, but less than once a day, while 29 women reported using cannabis at least once a day (coded 0/1/2 for analyses). Of these women, 20 reported using both tobacco and cannabis. To determine whether social disadvantage continued to predict residual birthweight even accounting for cannabis and tobacco use, we created a revised SEM model. We constructed an SEM that included this association, and which allowed us to both examine cannabis and tobacco use as part of the pathway by which *Social Advantage* might be linked to birthweight (e.g., an indirect pathway), versus a direct pathway from *Social Advantage* independent of cannabis/tobacco use. As shown in Table S2, cannabis and tobacco use were significantly and strongly associated with the *Social Advantage* latent factor, and more modestly associated with the *Psychosocial Distress* latent factor. The total effect for the relationship of *Social Advantage* to residual birthweight (combination of both direct and indirect pathways) remained highly significantly. Critically, the direct relationship of *Social Advantage* (i.e., independent of tobacco and cannabis use) to residual birthweight remained significant, while there was no direct relationship of cannabis or tobacco use to birthweight independent of *Social Advantage*. These data indicate that while cannabis and tobacco use are greater among those women experiencing greater social disadvantage and psychosocial distress, social advantage still predicts birthweight even accounting for this substance use.

Supplemental Table 1. Frequency timing and source of clinical and demographic variables

| **Variable** | **Frequency of measurement** | **Timing if singular** | **Composite value** | **Source** |
| --- | --- | --- | --- | --- |
| Income to Needs | Each trimester | NA | No | Self-reported |
| Insurance status | Singular | At enrollment | No | Medical Record |
| Maternal education | Singular | At enrollment | NA | Self-reported |
| ADI | Twice | At enrollment and third trimester | NA | Medical Record at enrollment and verified via self-report at third trimester |
| Healthy Eating Index | Singular | At third trimester or delivery | NA | DHQII self-reported |
| EPDS | Each trimester | NA | No | Self-reported |
| PSS | Each trimester | NA | No | Self-reported |
| STRAIN | Singular | At neonatal scan/1 yr. | No | Self-reported |
| Discrimination | Singular | At neonatal scan | No | Self-reported |
| Maternal age | Singular | At delivery | NA | Medical Record |
| Pre-pregnancy BMI | Singular | At enrollment | NA | Self-reported from Medical Record |
| MMR* | Preexisting conditions and over course of pregnancy | Enrollment to delivery | Yes 24 factors | Medical Record |
| Birthweight | Singular | At delivery | NA | Medical Record |
| Gestational age | Singular | At delivery | NA | Medical Record |

*Maternal co-morbidities included chronic congestive heart failure, congenital heart disease, pulmonary hypertension, chronic ischemic heart disease, cardiac valvular disease, pre-existing hypertension, gestational hypertension, chronic renal disease, preexisting diabetes mellitus, , asthma, severe pre-eclampsia, sickle cell disease, age ≥35, systemic lupus erythematousus, HIV virus, mild or unspecified pre-eclampsia, placental previa.

**Supplemental Table 2: Revised SEM Model Incorporating Cannabis and Tobacco Use**

| **Variable** | **Raw Estimate** | **STDY Estimate** | **Standard Error** | **Estimate/**  **Standard Error** | **p**  **(2-tailed)** | **Residual**  **Variance** |
| --- | --- | --- | --- | --- | --- | --- |
| **Indicators of Social Advantage** | | | | | | |
| Income/Needs 1^st^ Trimester | 0.364 | 0.904 | 0.019 | 48.087 | 0.000 | 0.182 |
| Income/Needs 2^nd^ Trimester | 0.371 | 0.913 | 0.019 | 48.864 | 0.000 | 0.167 |
| Income/Needs 3^rd^ Trimester | 0.370 | 0.912 | 0.017 | 54.230 | 0.000 | 0.168 |
| Area Deprivation Index | -17.727 | -0.717 | 0.029 | -24.417 | 0.000 | 0.486 |
| Health Insurance | 2.901 | 0.848 | 0.029 | 29.681 | 0.000 |  |
| Education | 2.834 | 0.842 | 0.023 | 35.880 | 0.000 |  |
| HEI-2016 Total Score | 3.766 | 0.382 | 0.054 | 7.140 | 0.000 | 0.854 |
| **Indicators of Psychosocial Distress** | | | | | | |
| EPDS 1^st^ Trimester | 3.642 | 0.749 | 0.033 | 22.899 | 0.000 | 0.440 |
| EPDS 2^nd^ Trimester | 3.832 | 0.771 | 0.030 | 25.474 | 0.000 | 0.406 |
| EPDS 3^rd^ Trimester | 3.384 | 0.706 | 0.051 | 13.795 | 0.000 | 0.502 |
| PSS 1^st^ Trimester | 5.717 | 0.772 | 0.029 | 26.443 | 0.000 | 0.404 |
| PSS 2^nd^ Trimester | 6.330 | 0.812 | 0.030 | 27.149 | 0.000 | 0.341 |
| PSS 3^rd^ Trimester | 5.344 | 0.709 | 0.039 | 18.406 | 0.000 | 0.497 |
| STRAIN - CT | 2.626 | 0.435 | 0.051 | 8.554 | 0.000 | 0.811 |
| STRAIN – WT SEV | 9.363 | 0.473 | 0.051 | 9.244 | 0.000 | 0.776 |
| Discrimination Survey | 0.523 | 0.235 | 0.059 | 4.016 | 0.000 | 0.945 |
| **Correlates of Social Advantage** | | | | | |  |
| Maternal Age at Delivery | 2.174 | 0.409 | 0.050 | 8.205 | 0.000 | 0.818 |
| Pre-Pregnancy BMI | -1.093 | -0.131 | 0.068 | -1.921 | 0.055 | 0.980 |
| Cannabis Use | -0.127 | 0.230 | 0.033 | -6.925 | 0.000 | 0.901 |
| Tobacco Use | -0.121 | 0.241 | 0.037 | -6.497 | 0.000 | 0.907 |
| **Correlates of Psychosocial Distress** | | | | | |  |
| Maternal Age at Delivery | -0.235 | -0.044 | 0.056 | -0.795 | 0.427 |  |
| Pre-Pregnancy BMI | 0.188 | 0.023 | 0.072 | 0.314 | 0.753 |  |
| Cannabis Use | 0.081 | 0.148 | 0.060 | 2.457 | 0.014 |  |
| Tobacco Use | 0.060 | 0.121 | 0.062 | 21.944 | 0.052 |  |
| **Predictors of Birthweight** | | | | | | 0.925 |
| Social Advantage | 0.008 | 0.138 | 0.070 | 1.983 | 0.047 |  |
| Psychosocial Distress | 0.001 | 0.008 | 0.055 | 0.154 | 0.878 |  |
| Maternal Medical Risk | 0.000 | -0.003 | 0.050 | -0.069 | 0.945 |  |
| Maternal Age at Delivery | 0.001 | 0.061 | 0.063 | 0.979 | 0.328 |  |
| Pre-Pregnancy BMI | 0.001 | 0.125 | 0.060 | 2.083 | 0.037 |  |
| Cannabis Use | -0.010 | -0.088 | 0.052 | -1.697 | 0.093 |  |
| Tobacco Use | -0.013 | -0.105 | 0.061 | 1.729 | 0.084 |  |
| **Direct and Indirect Predictors of Birthweight from Social Advantage** | | | | | |  |
| Social Advantage - Total | 0.012 | 0.192 | 0.057 | 3.370 | 0.001 |  |
| Social Advantage - Indirect | 0.003 | 0.054 | 0.038 | 1.412 | 0.158 |  |
| Maternal Age at Delivery | 0.002 | 0.025 | 0.026 | 0.957 | 0.338 |  |
| Pre-Pregnancy BMI | -0.001 | -0.016 | 0.012 | -1.395 | 0163 |  |
| Cannabis Use | 0.001 | 0.020 | 0.013 | 1.609 | 0.108 |  |
| Tobacco Use | 0.002 | 0.025 | 0.015 | 1.663 | 0.096 |  |
| Social Advantage - Direct | 0.008 | 0.138 | 0.070 | 1.983 | 0.047 |  |
| **Direct and Indirect Predictors of Birthweight from Psychological Distress** | | | | | |  |
| Psychological Distress - Total | -0.001 | -0.017 | 0.056 | -0.303 | 0.762 |  |
| Psychological Distress - Indirect | -0.002 | -0.026 | 0.017 | -1.489 | 0.136 |  |
| Maternal Age at Delivery | 0.000 | -0.003 | 0.004 | -0.628 | 0.530 |  |
| Pre-Pregnancy BMI | 0.000 | 0.003 | 0.009 | 0.300 | 0.764 |  |
| Cannabis Use | -0.001 | -0.013 | 0.009 | -1.418 | 0.156 |  |
| Tobacco Use | -0.001 | -0.013 | 0.010 | -1.334 | 0.182 |  |
| Psychological Distress - Direct | 0.001 | 0.008 | 0.055 | 0.154 | 0.878 |  |
| **Covariances** | | | | | | |
| **Computed from confirmatory factor analysis** | |  |  |  |  |  |
| Social Advantage w Psychosocial Distress | | -0.354 | 0.045 | -7.851 | 0.000 |  |
| **Inserted because of residual correlations** | |  |  |  |  |  |
| Income/Needs 1^st^ Trimester w 2^nd^ Trimester | | 0.736 | 0.059 | 12.573 | 0.000 |  |
| Income/Needs 1^st^ Trimester w 3^rd^ Trimester | | 0.680 | 0.062 | 11.025 | 0.000 |  |
| Income/Needs 2^nd^ Trimester w 3^rd^ Trimester | | 0.759 | 0.050 | 15.156 | 0.000 |  |
| STRAIN – STRAIN-CT w STRAIN-WTSEV | | 0.916 | 0.011 | 86.879 | 0.000 |  |
